# Genomic epidemiology reveals 2022 mpox epidemic in New York City governed by heavy-tailed sexual contact networks

**DOI:** 10.1101/2024.07.30.24311083

**Authors:** Jonathan E. Pekar, Yu Wang, Jade C. Wang, Yucai Shao, Faten Taki, Lisa A. Forgione, Helly Amin, Tyler Clabby, Kimberly Johnson, Lucia V. Torian, Sarah L. Braunstein, Preeti Pathela, Enoma Omoregie, Scott Hughes, Marc A. Suchard, Tetyana I. Vasylyeva, Philippe Lemey, Joel O. Wertheim

## Abstract

The global mpox epidemic in 2022 was likely caused by transmission of mpox virus (MPXV) through sexual contact networks, with New York City (NYC) experiencing the first and largest outbreak in the United States. By performing a phylogeographic and epidemiological analysis of MPXV, we identify at least 200 introductions of MPXV into NYC and 84 leading to onward transmission. Through a comparative analysis with human immunodeficiency virus (HIV) in NYC, we find that both MPXV and HIV genomic cluster sizes are best fit by scale-free distributions and that people in MPXV clusters are more likely to have previously received an HIV diagnosis (odds ratio=1.58; *p*=0.012) and be a member of a recently growing HIV transmission cluster, indicating overlapping sexual contact networks. We then model the transmission of MPXV through sexual contact networks and show that highly connected individuals would be disproportionately infected at the start of an epidemic, thereby likely resulting in the exhaustion of the most densely connected parts of the sexual network. This dynamic explains the rapid expansion and decline of the NYC outbreak, as well as the estimated cumulative incidence of less than 2% within high-risk populations. By synthesizing the genomic epidemiology of MPXV and HIV with epidemic modeling, we demonstrate that MPXV transmission dynamics can be understood by general principles of sexually transmitted pathogens.

## Introduction

The global mpox (previously referred to as monkeypox) epidemic was first identified in May 2022, starting with the diagnosis of a case in the United Kingdom, and swiftly followed by localized outbreaks across the world. Phylodynamic analyses have demonstrated that repeated cross-country transmissions of monkeypox virus (MPXV) seeded these outbreaks, made distinguishable due to an elevated evolutionary rate attributable to APOBEC3-associated hypermutation^1–6^. By the end of 2022, the global case count of this epidemic exceeded 80,000 and the United States was the country with the largest number of cases at 30,092^7^, with the earliest importations into the United States likely coming from Western Europe^4^. Within the United States, New York City (NYC) experienced the first and largest major outbreak with 3,821 cases diagnosed by the end of 2022^8,9^. NYC also conducted the first mpox virus vaccination campaign in the country, which was initiated in June 2022.

During the 2022 epidemic, MPXV is believed to have primarily spread through sexual contact networks among men who have sex with men (MSM)^10,11^, whereas previous outbreaks were the result of zoonotic and household transmissions^12^. Epidemic modeling suggests that the rapid global emergence, expansion, and decline of mpox during the 2022 epidemic could be explained by the transmission of MPXV through heavy-tailed sexual contact networks^13–16^, a heterogeneous network in which a subset of individuals have a disproportionately high number of contacts. Such networks account for the transmission patterns of other sexually transmitted pathogens, including human immunodeficiency virus type 1^17–20^ (HIV-1), hepatitis C virus^21^, and syphilis^22–26^. Phylodynamic analyses of sexually transmitted pathogens, including HIV, have shown that they recapitulate these heavy-tailed sexual contact networks^18–21^; however, these hypothesized dynamics have not yet been confirmed for MPXV by empirical analyses.

The mpox outbreak in NYC peaked shortly after the initiation of its vaccination campaign, but it is unclear whether the subsequent declining incidence of mpox was due to vaccination, behavioral changes, sexual contact network dynamics, or some combination thereof^27^. Long-term public health surveillance in NYC provides a distinct opportunity to examine whether the transmission dynamics of MPXV are consistent with those of other sexually transmitted infections, specifically HIV-1, for which molecular surveillance data have been collected for two decades and sexual transmission dynamics are well understood^17–20,28,29^. Here, we employed phylogeographic and epidemiological modeling approaches to better understand the factors governing the trajectory of the mpox outbreak in NYC in the context of the global epidemic and the intersection between the MPXV and HIV transmission networks.

## Results

### Emergence of mpox in NYC

The mpox outbreak in NYC rapidly grew in the spring of 2022 (Fig. 1a), while the rest of the United States did not experience a significant increase in cases until the end of July. Mpox cases in NYC peaked in late July, approximately a month before the peaks in the rest of the United States and globally. A substantially higher percentage of diagnosed mpox infections were sequenced in NYC through April 2023 (19.7%; *n*=760) than in either the rest of the United States (4.6%; n=1221) or rest of the world (4.1%, n=2338), allowing for a fine-grained phylogeographic analysis of the MPXV outbreak in NYC. Over 99% of the MPXV genomes from NYC were produced by the NYC Department of Health and Mental Hygiene (DOHMH) Public Health Laboratory.

**Figure 1.**
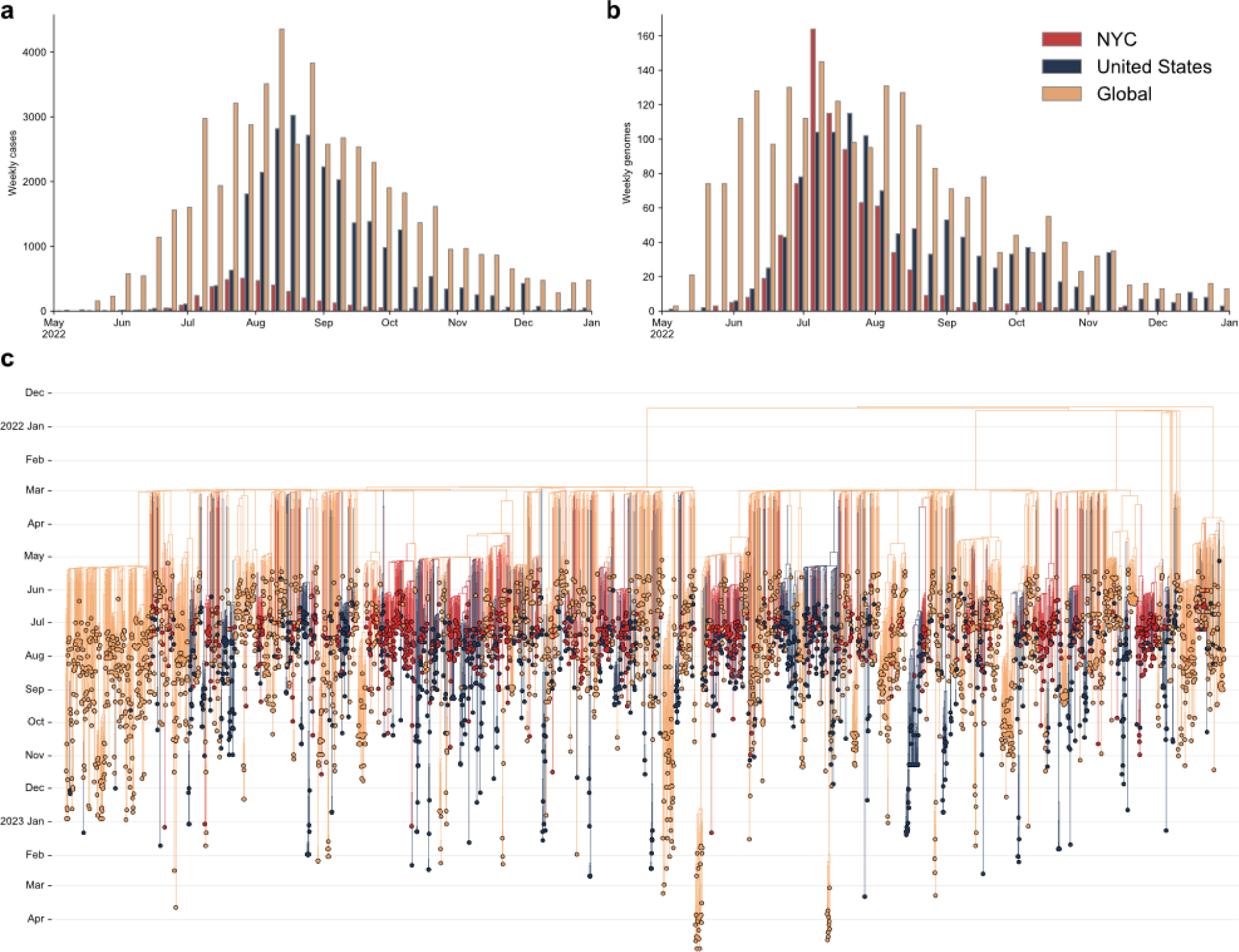
Emergence of the 2022 mpox epidemic in New York City (NYC). **a**, The number of cases of mpox per week in NYC, the United States, and globally, where ‘United States’ refers to all of the United States excluding NYC and ‘global’ refers to all countries other than the United States. **b,** The number of sequenced genomes per week in NYC, the United States, and globally. **c,** Time-calibrated phylogenetic tree of MPXV.

To characterize the introductions and emergence of MPXV in NYC, we conducted a Bayesian phylogeographic analysis of MPXV sampled from April 2022 through April 2023, with 757 high-quality genomes from NYC (756 sequenced by NYC Public Health Laboratory [PHL]), 1127 from the rest of the United States, and 2160 from the rest of the world^30^. We found that MPXV was likely cryptically circulating in NYC and the rest of the United States for 1–2 months before the earliest cases of the 2022 mpox outbreak were identified on 19 May and 17 May in NYC and the broader United States, respectively (Fig. 1c), in line with previous results^4^. We inferred that global importations of MPXV into NYC started in April, with a peak in mid-June (Fig. 2a), and identified 174 global importations into NYC in total (posterior median, 95% highest posterior density [HPD]: 160–191). Based on the available sampling, there were substantially fewer importation events into NYC from the broader United States (posterior median: 59; 95% HPD: 43–75), with both importations from and exportations into the United States peaking in late June to early July (Fig. 2a). However, there were many exportations of MPXV from NYC to the rest of the United States (posterior median: 170; 95% HPD: 143–193; Fig. 2b).

**Figure 2.**
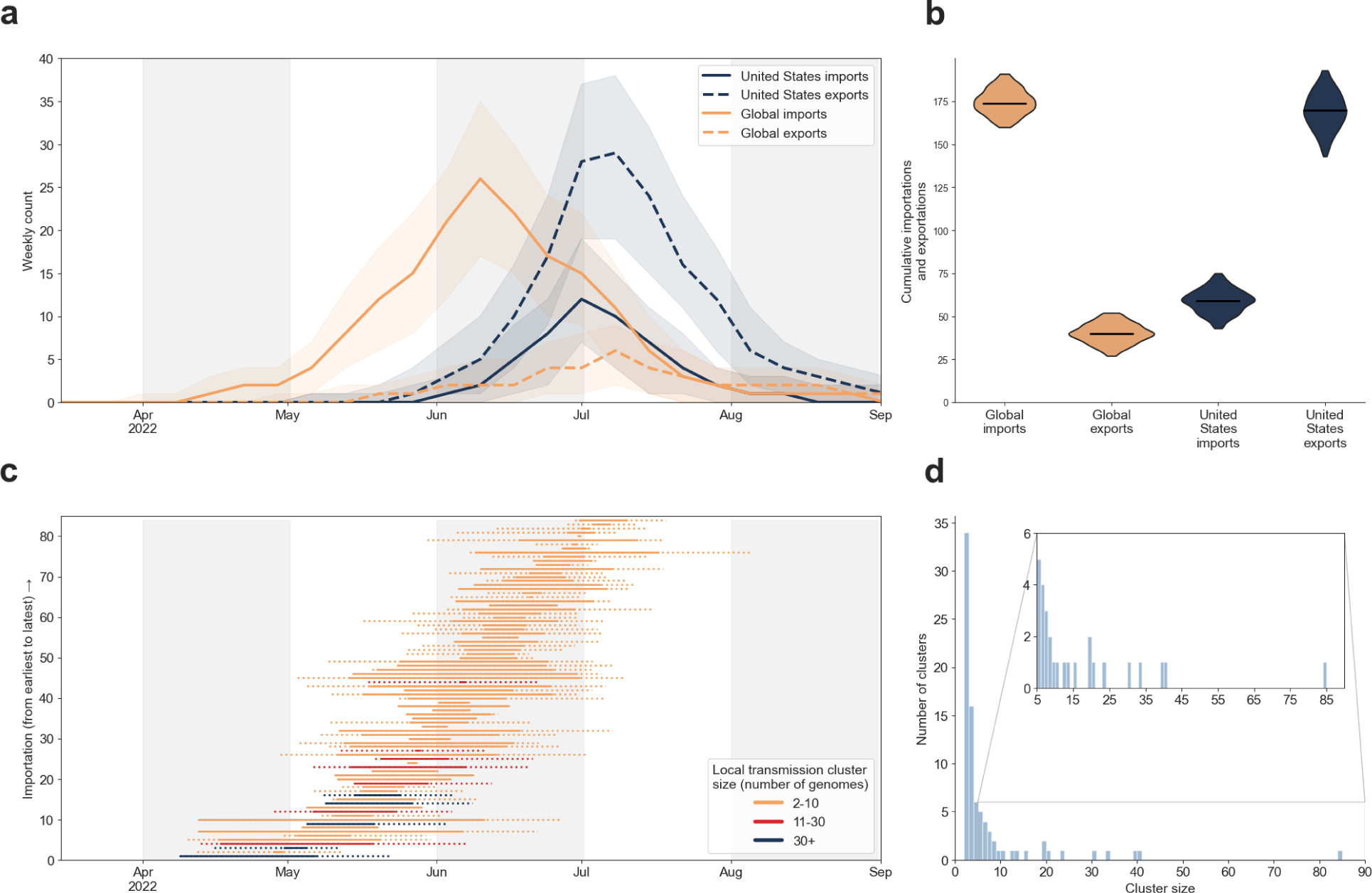
Importations and exportations of MPXV in NYC. **a,** The estimated number of MPXV importations into and exportations out of NYC per week. Shaded areas show 95% highest posterior densities (HPDs) of the estimates. **b,** Cumulative number of MPXV importations into and exportations out of NYC. The violins represent the 95% HPDs and the black bars indicate the median values. **c,** Estimated importation times for the 84 inferred MPXV NYC transmission clusters. The left limit of the solid line is the median time of the parent node of the MRCA of each transmission cluster and the right limit is the median tMRCA of the transmission cluster. The left limit of the dotted line is the lower bound of the 95% HPD of the time of the parent node and the right limit is the upper bound of the 95% HPD of the tMRCA. **d,** Distribution of MPXV transmission cluster sizes in NYC.

Global importations seeded the start of the epidemic in the rest of the United States (Extended Data Fig. 1). Then, starting in July, most importations into the broader United States came from NYC. By the end of 2022, there were nearly as many importations from NYC into other areas of the United States as from the rest of the world.

### MPXV transmission clusters

Our phylogeographic analysis allows for the identification of NYC transmission clusters: distinct importations of MPXV into NYC that result in identifiable onward transmission within NYC (*i.e.*, at least two individuals with sequenced MPXV genomes). We inferred 84 clusters in total, and the importations of MPXV into NYC that led to the largest transmission clusters are estimated to have occurred in late April or early-to-mid May 2022 (Fig. 2c). However, most imports, especially later in the outbreak, led to singletons: introductions characterized by a single viral genome without any identifiable onward transmission in NYC (Extended Data Fig. 2).

We assessed the association between demographic attributes and MXPV transmission risk reported to public health surveillance and the frequency of membership in MPXV transmission clusters among 703 individuals (703/756; 93.0%) for whom this information was reported. This population mostly comprised MSM (Table 1), and they resided in all five NYC boroughs. We observed increased odds of membership in an MPXV transmission cluster for people residing in Brooklyn (*p*=0.006; Table 1) and individuals identifying as Black/African American (*p*=0.038). There were decreased odds of MPXV clustering for individuals identifying as White (*p*=0.005) and those with recent foreign travel history (*p*=0.001). An explanation for this inferred negative association between clustering and foreign travel is found in our definition of clusters as unique introductions into NYC. Although each transmission cluster must have, by definition, been initiated by someone with recent travel history, introductions that did not lead to onward transmission would include only an individual with foreign travel history. (Fig. 2d, Extended Data Fig. 2). Membership in a cluster did not vary significantly by gender, sexual orientation, age, or vaccination status (Table 1).

**Table 1.**
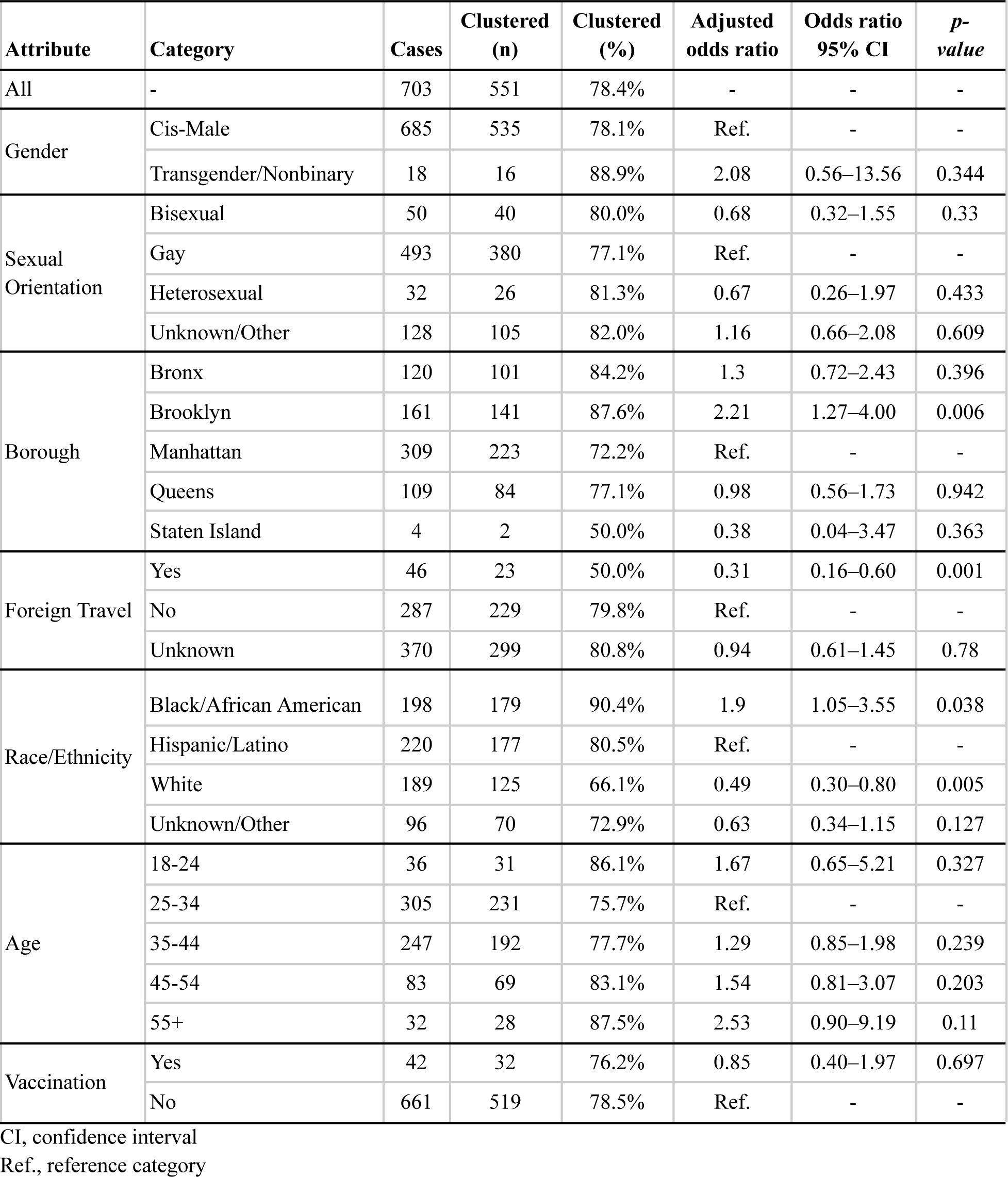
Attributes associated with membership in MPXV clusters using multivariate logistic regression model.

### Associations between MPXV and HIV transmission clusters

The distribution of MPXV transmission cluster sizes is highly right-skewed (Fig. 2d), matching previously described patterns of HIV transmission cluster size distributions among MSM^18–20^. To better characterize these MPXV transmission cluster sizes, we fit a set of distributions to them and found that scale-free distributions provided the best fit (AIC < 550; Table 2). To validate the fit of these distributions, we simulated transmission cluster sizes from them and calculated how similar the simulated cluster sizes were to the inferred cluster sizes. Scale-free distributions consistently fit the inferred MPXV transmission cluster sizes better than distributions that were not scale-free, with a Yule-Simon distribution being the best fit (Kullback–Leibler [KL] divergence = 0.336).

**Table 2.** Distributions fitted to MPXV and HIV transmission clusters.

| Distribution | MPXV |  | HIV |  |
| --- | --- | --- | --- | --- |
|  | AIC | KL Divergence | AIC | KL Divergence |
| Binomial | 1409.716 | 1.737 | 38,082.988 | 1.922 |
| Exponential | 582.725 | 0.757 | 20,822.706 | 0.716 |
| Negative Binomial | 602.169 | 0.517 | 21,237.044 | 0.471 |
| Power Law<sup>1</sup> | 461.878 | 0.477 | 18,483.141 | 0.413 |
| Pareto<sup>1</sup> | 521.209 | 0.429 | 21,929.536 | 0.339 |
| Yule Simon<sup>1</sup> | 507.858 | 0.327 | 21,254.079 | 0.247 |

<sup>1</sup>Scale-free distributions.

We then compared the cluster size distributions of MPXV and HIV in NYC. We first inferred transmission clusters from HIV partial*-pol* sequences sampled since 2001 from 76,910 people living with HIV (PLWH) in NYC using HIV-TRACE, identifying 4259 HIV transmission clusters comprising 17,456 individuals. We found 416 HIV transmission clusters that grew by at least one member in 2021 or 2022, comprising 4097 people. Of these people in growing clusters, 1711 (40.2%) had a virus with a recent genetic link to another person with an HIV diagnosis in 2021 or 2022. Fitting distributions to the HIV transmission cluster sizes, we found that the HIV transmission cluster sizes in NYC were also right-skewed (Extended Data Fig. 3) and best fit by scale-free distributions (Table 2). Both MPXV and HIV transmission clusters in NYC follow scale-free distributions, recapitulating transmission across scale-free sexual transmission networks with heavy-tailed partnership distributions^31^ and suggesting their transmission dynamics likely emerged from similar forms of transmission.

We queried the HIV surveillance database in NYC to determine if persons living with HIV and in the HIV transmission network were disproportionately represented in MPXV clusters (Table 3). A substantial proportion of the individuals with an mpox diagnosis had a reported HIV diagnosis (n=328/756; 43.4%), and we found a significant association between a reported HIV diagnosis and membership within MPXV transmission clusters (logistic regression; odds ratio=1.58; *p*=0.012). The majority (310/328, 94.5%) of people with both an HIV and mpox diagnosis received their HIV diagnosis prior to 2022, highlighting the different time frame in which these viruses spread among the same people (Extended Data Fig. 4). Individuals who were clustered in both the MXPV network and the HIV network did not share connections in their networks, suggesting that the contact networks through which MPXV and HIV were non-overlapping, in line with the different time periods in which individuals received MPXV and HIV diagnoses.

**Table 3.**
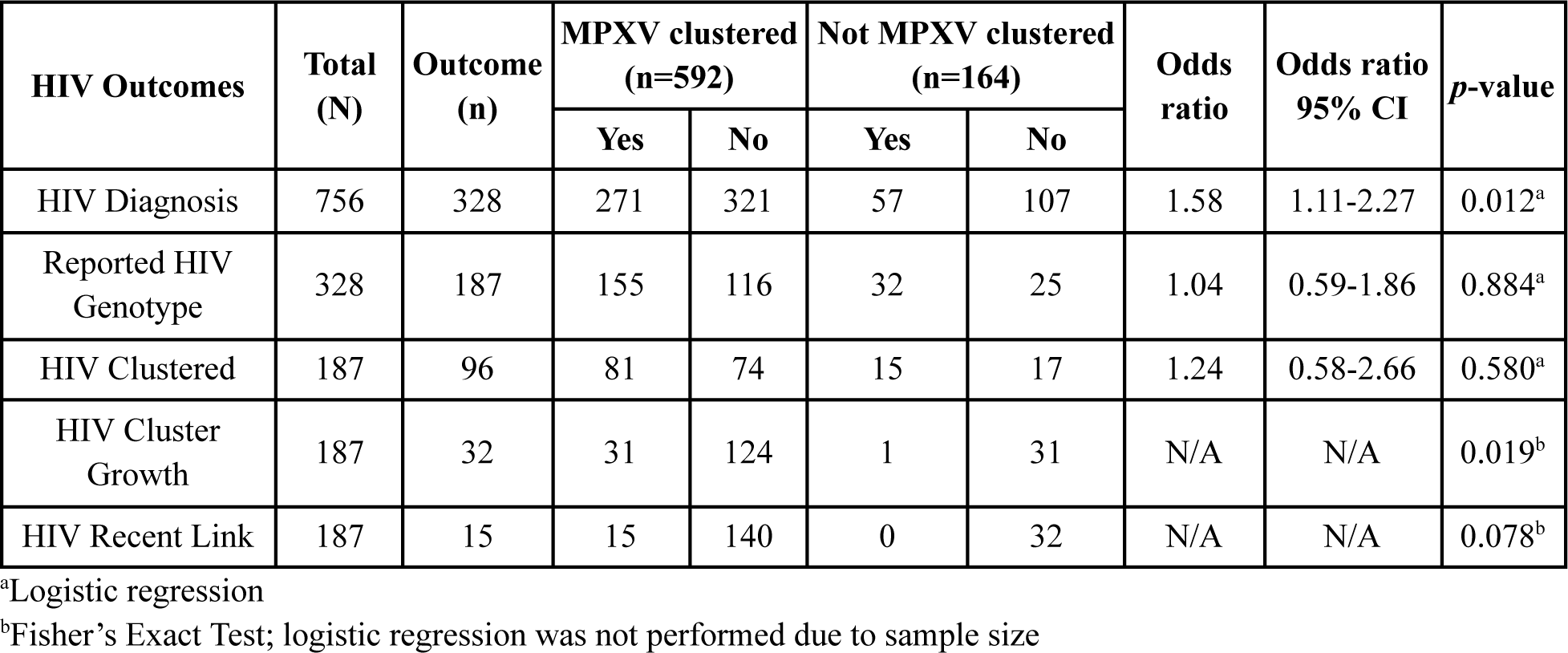
MPXV and HIV transmission lineage and cluster matching. Breakdown of 756 individuals with reported MPXV genomes from PHL included in the phylogeographic analysis based on the clustering profile, association with the HIV transmission network, and related information from HIV surveillance database.

| HIV Outcomes | Total (N) | Outcome (n) | MPXV clustered (n=592) |  | Not MPXV clustered (n=164) |  | Odds ratio | Odds ratio 95% CI | p-value |
| --- | --- | --- | --- | --- | --- | --- | --- | --- | --- |
|  |  |  | Yes | No | Yes | No |  |  |  |
| HIV Diagnosis | 756 | 328 | 271 | 321 | 57 | 107 | 1.58 | 1.11-2.27 | 0.012<sup>a</sup> |
| Reported HIV Genotype | 328 | 187 | 155 | 116 | 32 | 25 | 1.04 | 0.59-1.86 | 0.884<sup>a</sup> |
| HIV Clustered | 187 | 96 | 81 | 74 | 15 | 17 | 1.24 | 0.58-2.66 | 0.580<sup>a</sup> |
| HIV Cluster Growth | 187 | 32 | 31 | 124 | 1 | 31 | N/A | N/A | 0.019<sup>b</sup> |
| HIV Recent Link | 187 | 15 | 15 | 140 | 0 | 32 | N/A | N/A | 0.078<sup>b</sup> |

<sup>a</sup>Logistic regression
<sup>b</sup>Fisher's Exact Test; logistic regression was not performed due to sample size

Clustering in the HIV network is associated with more densely connected sexual networks and rapid viral transmission^17,28,32^. We did not observe an association between mpox diagnoses with reporting of HIV sequences or clustering in the HIV transmission network. However, the frequency with which people with an mpox diagnosis clustered in the HIV network (n=96/187; 51.3%) is substantially higher than would be expected; resampling the HIV network matching transmission risk and diagnosis year of people with an mpox diagnosis, one would expect only 67 (35.8%; 95% range: 55–80) individuals clustered in the HIV network. These results indicate that people with both HIV and mpox diagnoses are part of more densely connected sexual networks.

This association of mpox cases with active sexual networks is further demonstrated by their increased occurrence in recently growing HIV transmission clusters (Fisher’s exact test; *p*=0.019). Nearly all individuals with an mpox diagnosis and membership in a growing HIV cluster (i.e., ≥1 new member diagnosed in 2021-2022) were also part of an MPXV cluster (n=31/32). Further, all 15 individuals with an mpox diagnosis and a genetic link to someone diagnosed with HIV in 2021 or 2022 in the HIV transmission network belonged to an MPXV cluster, although the small dataset offers limited statistical power (Table 3).

### Decline of the NYC mpox outbreak

To quantify when and why the 2022 mpox outbreak in NYC began its decline, we inferred the time-varying reproduction number (also known as the effective reproduction number, R_e_) using case counts, generation time, and incubation period. The epidemic in NYC reached its peak at the end of July 2022 (Fig. 1a), before most of the first vaccine doses of a two-dose vaccine series were administered (Fig. 3a). We found that R_e_ peaked in late June and began decreasing before fewer than 1000 doses of the first vaccine dose were administered (*i.e.*, <1% of the 102,183 first vaccine doses delivered in NYC in 2022) (Fig. 3). Further, R_e_ decreased to below one by August 2022, before 200 doses of the second vaccine dose were administered (*i.e.*, <1% of the 52,374 second vaccine doses delivered in NYC in 2022).

**Figure 3.**
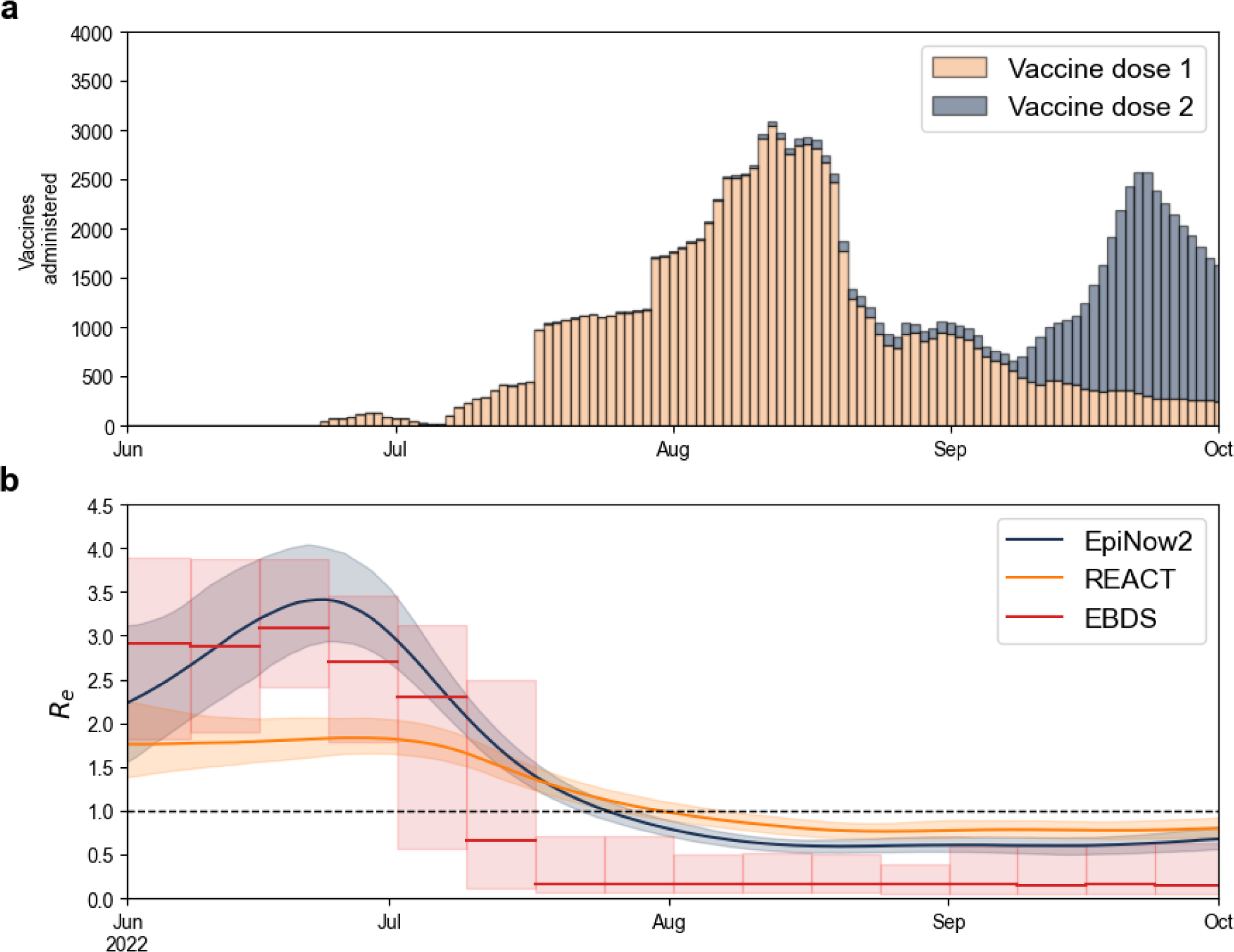
Vaccine administration and effective reproduction number over time. **a**, Number of mpox vaccines administered in mid-late 2022. **b**, Inferred effective reproduction number (R_e_) of MPXV based on the case data whilst incorporating generation time and incubation period into the inference (EpiNow2, blue), solely on the case counts (REACT, yellow), or MPXV genomes in a phylodynamic approach using an episodic birth-death-sampling model (EBDS; red). Dashed line indicates inflection point (*i.e.*, when the epidemic is in decline; R_e_=1). Lines and shaded regions represent the median and 95% confidence interval, respectively. The EBDS model assumes week-long intervals with a uniform R_e_ per interval.

Generation time and incubation period can vary throughout an epidemic^33^, thereby resulting in model misspecification. Therefore, we also inferred R_e_ using solely case counts and found that R_e_ could have dropped below 1 up to one week later (Fig. 3b), but still before a moderate proportion of the second vaccine was administered.

The local transmission clusters identified through our phylogeographic inference provide an alternative approach to measure changes in R_e_ through the NYC mpox epidemic that relies on genomics and inferred local transmission. We constructed an episodic birth-death-sampling (EBDS) model in a Bayesian framework that jointly analyzed all imported NYC clusters with at least 10 genomes (n=13). We found that R_e_ decreased below 1 by mid-July, approximately 1–2 weeks before the dates inferred when primarily using case data, in line with many of the introductions of MPXV into NYC after June not having identifiable onward transmission (Extended Data Fig. 2).

To confirm these results, we inferred the growth rate of mpox (Extended Data Fig. 5), as growth rate has been shown to be less sensitive than R_e_ to model misspecification^34^. We found that the growth rate also began its decline by the end of June, when fewer than 1000 doses of the first vaccine dose had been administered, and decreased below zero by August, approximately at the same time as the peak of the outbreak. The growth rate inference was consistent both with and without the generation time and incubation period parameterization. When focusing on local transmission clusters, we again found that the growth rate decreased below zero approximately 1–2 weeks prior to the inferred dates when using the other approaches.

### Heavy-tailed sexual contact network drives outbreak dynamics

To determine whether a heavy-tailed sexual contact network could explain the decline of the outbreak before there was substantial infection- or vaccine-derived immunity, we simulated mpox epidemics based on MSM sexual contact networks, under varying assumptions for the infectious period and the risk of transmission between sexual partners (secondary attack rate [SAR] during the infectious period). By the time R_e_ dropped below 1 in our simulations, we estimated a cumulative incidence among MSM of below 2% when assuming an SAR of 0.1 (Extended Data Fig. 6a); a higher SAR produces slightly higher average cumulative incidence, but with a larger range (Extended Data Fig. 6b). Assuming an MSM population in NYC of approximately 235,000 (see Methods), we observed an empirical cumulative incidence of 0.52–0.94% (n*=*1230–2217) at the time R_e_ dropped below 1 (Fig. 3). If we conservatively assume a 60% mpox case ascertainment rate^4,35^, there would have been an empirical cumulative incidence of 0.87–1.57% at this time. These results are broadly consistent with our simulations, suggesting that the dynamics of the mpox outbreak in NYC were likely governed by a heavy-tailed sexual network, with an SAR likely between 0.1 and 0.4 (Extended Data Fig. 6).

From these epidemic simulations, we found that the most highly connected individuals (i.e., people with more sexual partners than at least 99% of the sexual contact network) become infected early in the epidemic and nearly 40% of these individuals were infected by the time the empirical R_e_ dipped below 1 (Fig. 4a). This dynamic results in over 25% of all infections comprising highly connected individuals by the time R_e_ reached 1 (Fig. 4b); across varying connectivity thresholds, a disproportionate number of the cumulative cases are attributed to the highly connected individuals. The rapid infection of highly connected individuals in the sexual network at the start of the epidemic suggests their connectivity was a major contributor to the initially high R_e_ and that the early saturation of densely connected parts of the sexual network led to the epidemic ending with low cumulative incidence.

**Figure 4.**
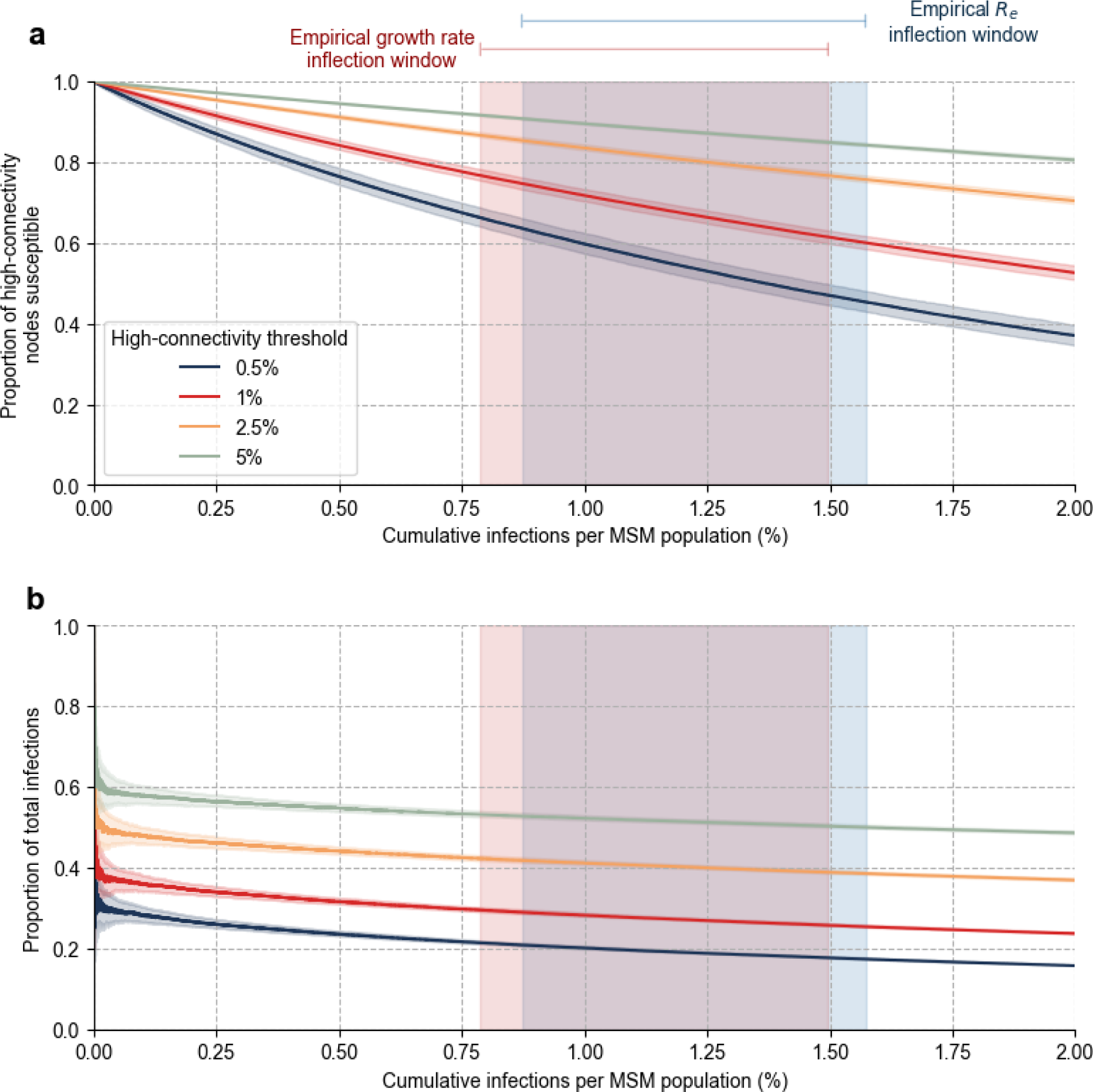
Infection of highly connected individuals. **a,** Proportion of highly connected susceptible individuals remaining after observing given cumulative cases. **b,** The proportion of cumulative infections made up by infected highly connected individuals. The high-connectivity threshold refers to the top percentile of network connectivity. Lines and shaded regions represent the median and 95% range, respectively. The red and blue rectangles represent when the inferred growth rate and *R_e_* reached their inflection points based on the empirical data. The left and right limits of the rectangles are, respectively, the lowest lower limit and the highest upper limit across the 95% confidence intervals of the methods based on case counts detailed in Figure 3 and Extended Data Figure 5.

## Discussion

We investigated the introductions and patterns of MPXV transmission clusters in NYC. Long-standing molecular surveillance in NYC presented an opportunity to investigate the associations between MPXV and HIV transmission networks and whether MPXV recapitulated the dynamics of a sexually transmitted pathogen like HIV. We found that earlier introductions of MPXV into NYC resulted in, on average, larger transmission clusters than later introductions, following model-based expectations^36^, and that the largest clusters were likely seeded before the first dozen cases were identified in NYC. These clusters were best described by a scale-free distribution, matching the cluster size distribution of HIV, and membership in these clusters was associated with HIV status and membership in growing HIV clusters. Our modeling subsequently showed that transmission through a heavy-tailed sexual contact network could explain the epidemic trends observed in NYC, regardless of changes in behavioral patterns or vaccine-induced immunity.

Sexual transmission networks, particularly among MSM, are characterized by heavy-tailed degree centrality distributions^13,37,38^, which are also observed in HIV transmission networks^19,20^. Considering the concordance between HIV and MPXV transmission networks and the similarities in their cluster size distributions, the distribution of contacts in the MPXV sexual network suggests a heavy-tailed partnership distribution as well. The extent to which these MPXV and HIV cluster-size distributions truly represent scale-free contact networks is a matter of ongoing debate^39,40^. However, cluster size distributions depend on the structure of the contact network, and heavy-tailed degree distributions are known to produce power-law cluster size distributions^31^.

We found that the 2022 mpox outbreak in NYC was consistent with the rapid and early infection of highly connected individuals in the sexual contact networks. Although these individuals make up a small proportion of the overall contact network, their immunity, whether from vaccination or natural infection, would disrupt chains of transmission. The decrease in transmission cluster sizes as the outbreak progressed is therefore likely the result of infection-derived immunity of these highly connected individuals^14^. It has similarly been suggested that the infection of highly connected individuals early in an epidemic could result in a decrease in frequency of exporting MPXV to other areas^14^. Our results corroborate this hypothesized pattern, with exportations of MPXV from NYC to the rest of the United States nearing zero before exportations from the rest of the world.

Contact network heterogeneity, changes in behavior, and vaccines could each individually impact MPXV transmission dynamics. Although there were undoubtedly changes in sexual behavior after a global mpox epidemic was declared^41,42^, changes in behavior at the peak of the epidemic were likely not the primary cause for its decline in NYC^13,14,16^. Even though the mpox vaccination campaign does not appear to be responsible for slowing and ultimately curbing the 2022 epidemic, vaccination—even a single dose—likely prevented many individuals from infection and severe disease^43–45^. Importantly, the lack of a resurgent mpox outbreak in NYC in the subsequent year, despite the continued circulation of MPXV among MSM^7^, suggests that vaccination is likely playing a role alongside natural (infection-induced) immunity in preventing renewed community transmission. Further, although NYC was able to rapidly and effectively distribute mpox vaccines to those at greatest risk of infection, the rise and fall of the epidemic appears to have outpaced the delivery of the vaccine, with natural immunity being the main contributor to the decline in NYC.

These results should be interpreted in the context of several limitations. First, uneven sampling and sequencing coverage could bias the inference of viral importations and onward transmission. Specifically, genomic sampling peaked in NYC before cases (Fig. 1a, b); however, considering NYC has the highest proportion of cases sequenced, it is unlikely that we overestimated the number of introductions of MPXV into NYC. Nearly all of the sequenced cases were from sexual health clinics, which, although not necessarily representative of citywide cases, are likely a sentinel population with which to understand sexually transmitted pathogens in NYC. Second, generation times and serial intervals of MPXV likely varied over the course of the epidemic, as evidenced by different estimates across regions and timeframes^46–49^. We therefore inferred the growth rate in addition to the effective reproduction number and used multiple approaches for the inference of each, arriving at consistent results for when the outbreak ended. Lastly, for our simulations, we restricted the population to MSM, assumed the sexual partnership distribution estimated for the MSM population in the United Kingdom was a sufficient proxy for that in NYC, and did not account for network structure or changes in behavior. However, heterosexual transmission of MPXV has not been shown to be a major factor in the mpox epidemic, and a more heterogeneous population than the one modeled or substantive shifts in behavior could result in an epidemic that could end with even lower cumulative incidence.

The transmission dynamics of the 2022 mpox outbreak in NYC was characterized by rapid spread through heavy-tailed sexual contact networks, which brought about the end of the epidemic when an estimated less than 2% of the at-risk MSM population had been infected. Although the World Health Organization declared an end to mpox as a public health emergency of international concern in May 2023, there have continued to be mpox cases both in NYC and elsewhere^50^, highlighting the potential for resurgence. Considering the dense connectivity of the sexual networks along with MPXV spreads, it is critical to develop targeted and rapid vaccination strategies not only in the event of future mpox outbreaks, but also as a preventative measure.

**Extended Data Figure 1.**
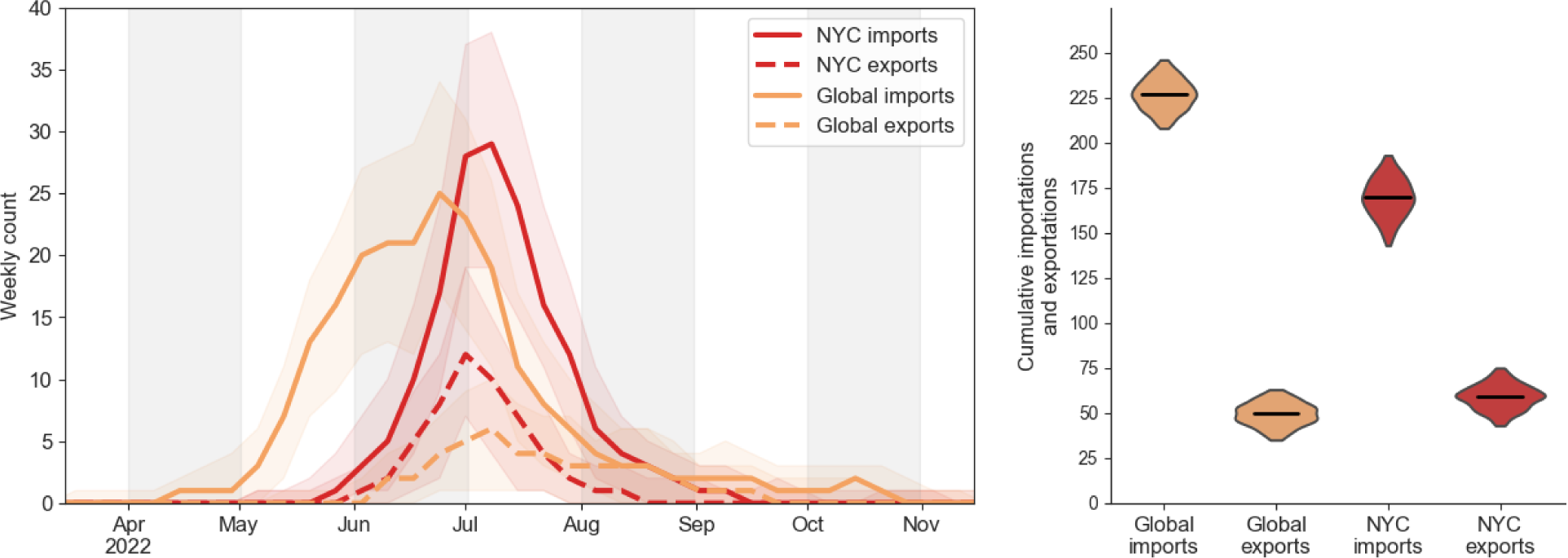
Importations and exportations of MPXV in the United States. **a,** The estimated number of MPXV importations into and exportations out of the United States per week. Shaded areas show 95% highest posterior densities (HPDs) of the estimates. **b,** Cumulative number of MPXV importations into and exportations out of the United States. The violins represent the 95% HPDs and the black bars indicate the median values.

**Extended Data Figure 2.**
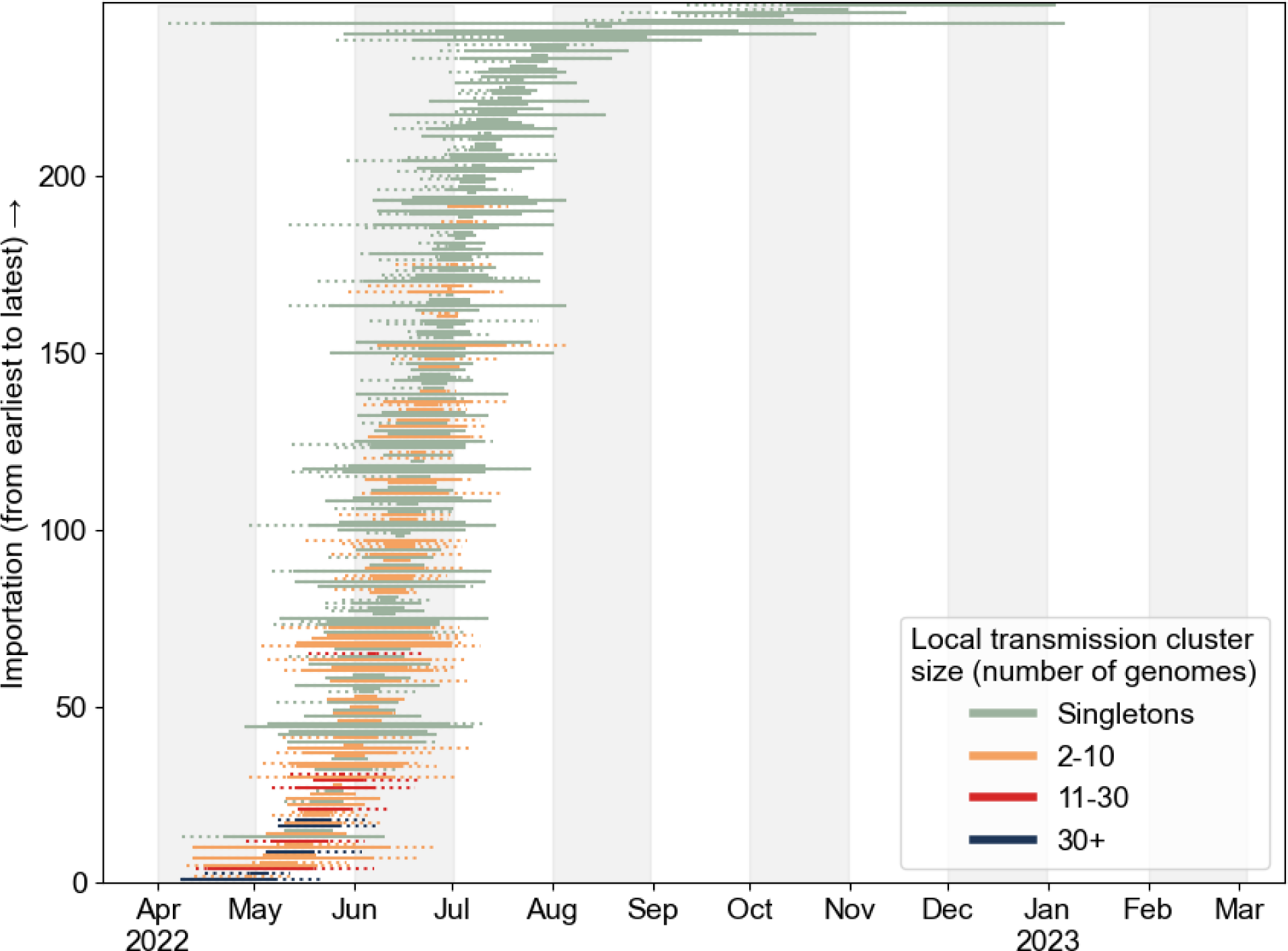
Estimated importation times for NYC transmission clusters, including singletons. The left limit of the solid line is the median time of the parent node of the MRCA of each transmission cluster and the right limit is the median tMRCA of the transmission cluster. The left limit of the dotted line is the lower bound of the 95% HPD of the time of the parent node and the right limit is the upper bound of the 95% HPD of the tMRCA. For singletons, the right limit of the solid line is the date of sampling, and there is no dotted line extending rightward.

**Extended Data Figure 3.**
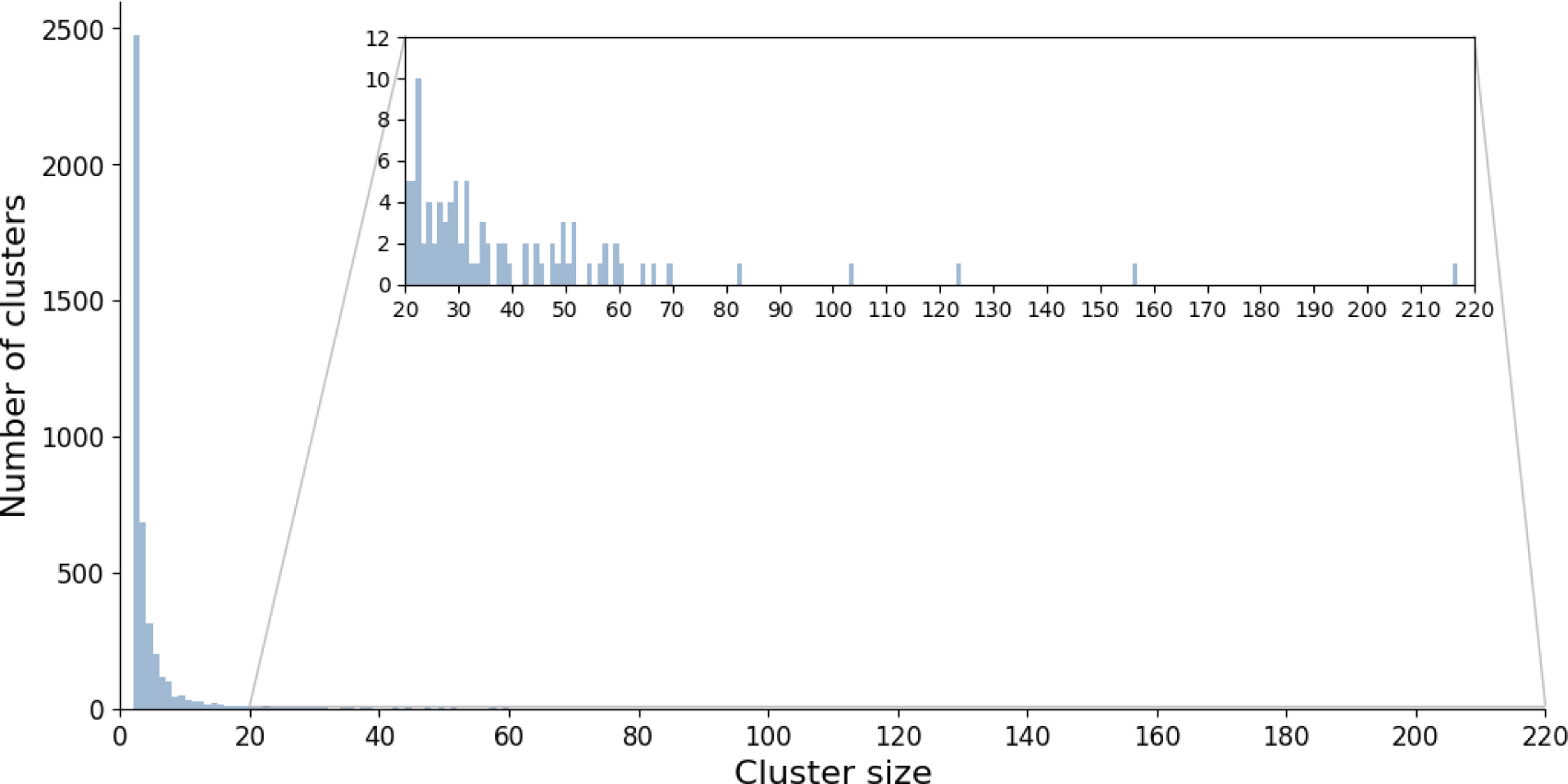
HIV transmission cluster sizes. Inset shows frequency of cluster sizes of at least 20.

**Extended Data Figure 4.**
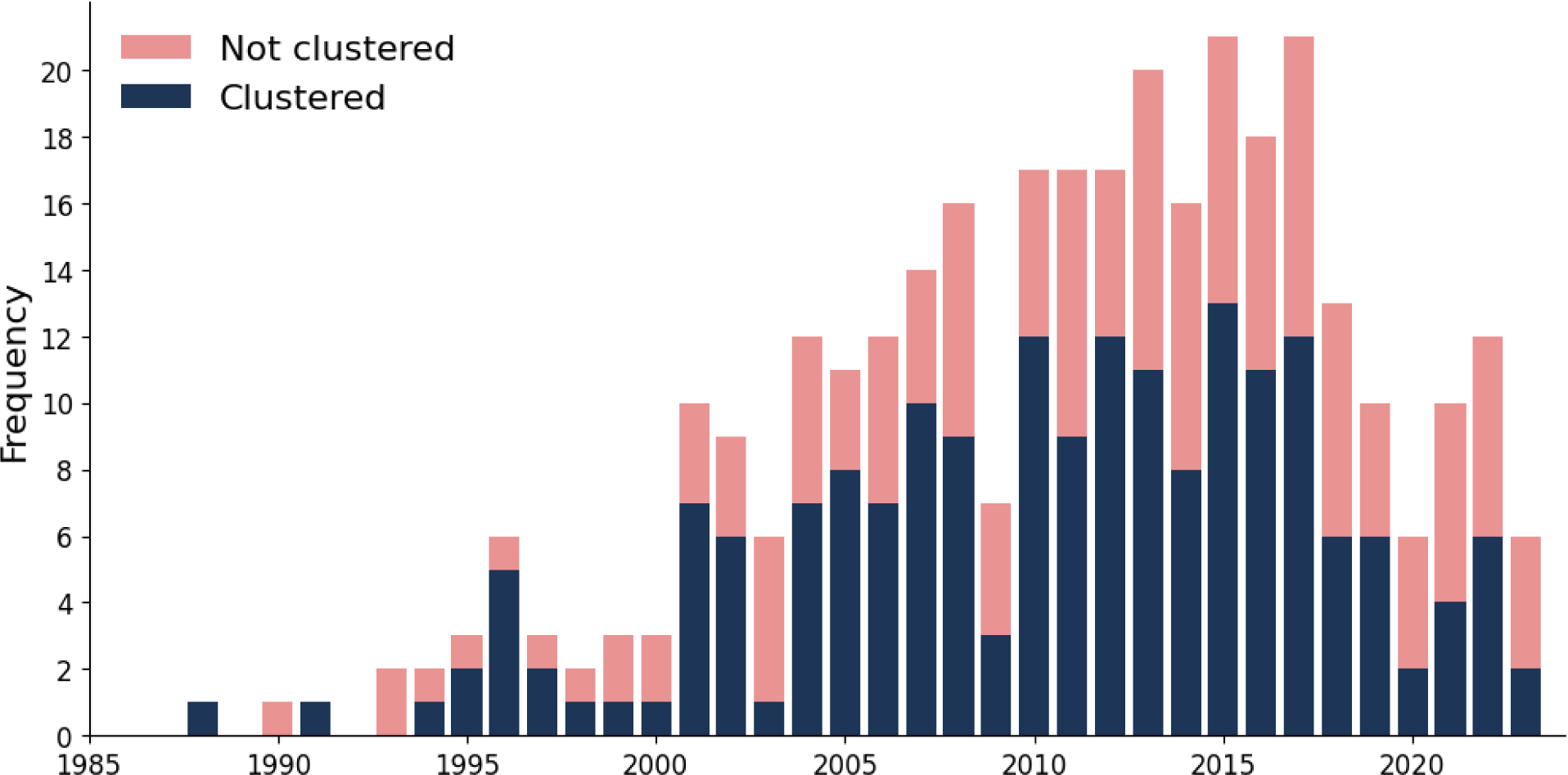
Year of HIV diagnosis for individuals diagnosed with mpox and living with HIV. The bars are colored by whether the individuals are members of mpox clusters.

**Extended Data Figure 5.**
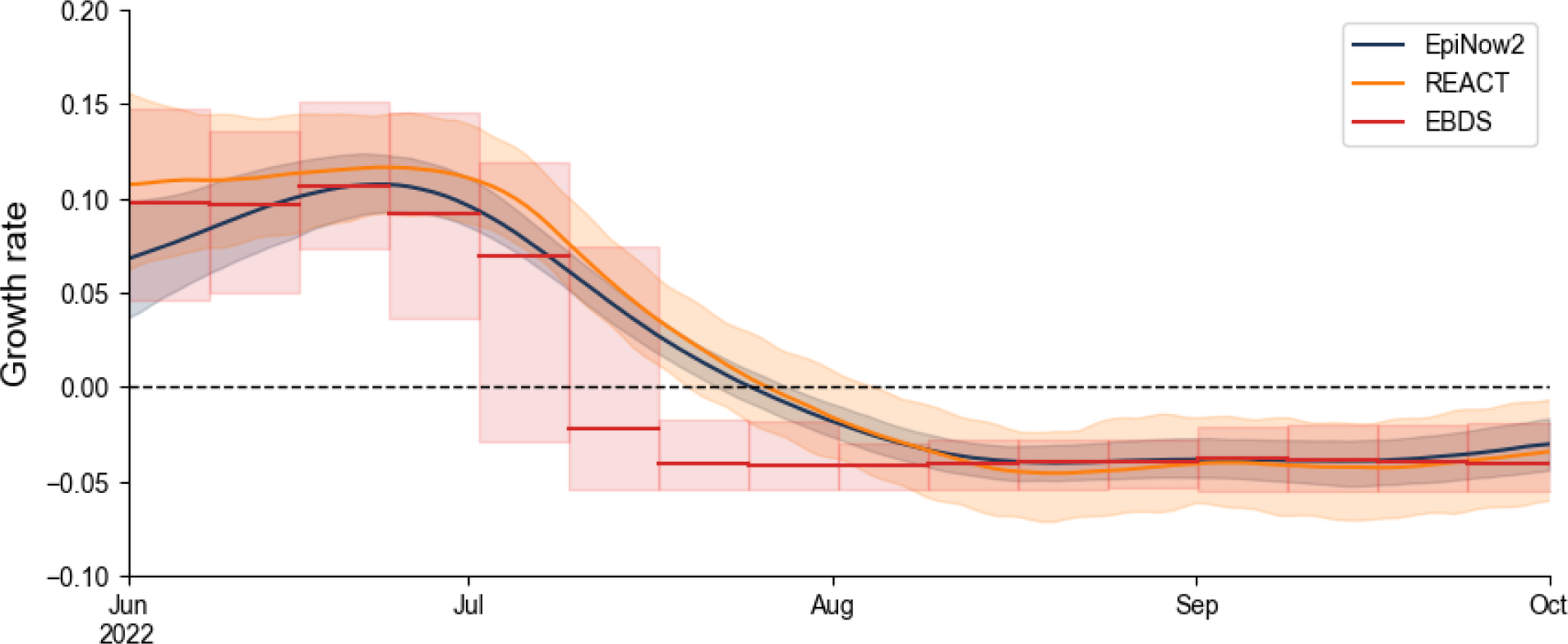
Effective reproduction number and growth rate of MPXV. Inferred growth rate of MPXV based on the case data whilst incorporating generation time and incubation period into the inference (EpiNow2, blue), solely on the case counts (REACT, yellow), or MPXV genomes in a phylodynamic approach using an episodic birth-death-sampling model (EBDS; red). Dashed line indicates inflection point (*i.e.*, when the epidemic is in decline; growth rate=0). Lines and shaded regions represent the median and 95% confidence interval, respectively. The EBDS model assumes week-long intervals with a uniform growth rate per interval.

**Extended Data Figure 6.**
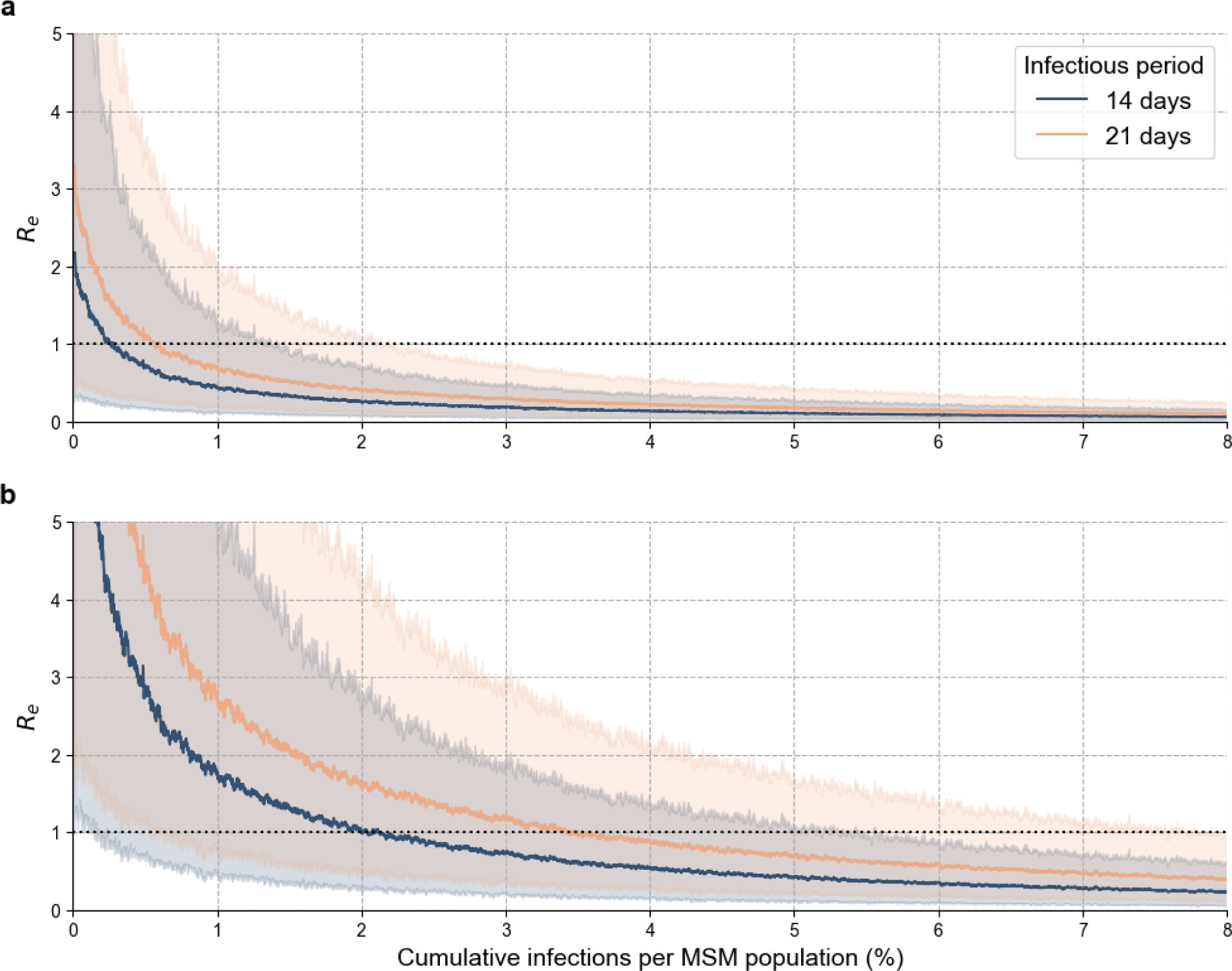
Effective reproduction number and depletion of highly connected susceptibles of simulated MPXV epidemics. The effective reproduction number (R_e_) after observing given cumulative infections with varying infectious periods and an SAR of (**a**) 0.1 and (**b**) 0.4. Lines and shaded regions represent the median and 95% range, respectively.

## Methods

### Incidence and vaccine data

The number of reported mpox cases per country were downloaded from Our World in Data (OWID; https://ourworldindata.org/; last accessed on 6 February 2024).

The number of reported mpox cases and vaccines administered in New York City (NYC) were generated by the NYC Department of Health and Mental Hygiene (DOHMH) (https://github.com/nychealth/monkeypox-data and https://github.com/nychealth/mpv-vaccine-data?tab=readme-ov-file, respectively; last accessed on 6 February 2024).

### Genomic data and initial processing

We generated 1,144 MPXV sequences from 759 individuals as part of public health surveillance by the NYC DOHMH Public Health Laboratory (PHL), which are available for download on NCBI as part of BioProject PRJNA949682.

We downloaded all available MPXV sequences from GISAID (last accessed 23 May 2023) while excluding the NYC sequences matching ours from NCBI. From these genomes, we subsequently excluded any genomes sampled before 28 April 2022 or after April 2023 and those that did not belong to the Clade II, B.1 lineage, resulting in 1 additional genome from NYC, 1221 from the rest of the United States, and 2338 from the rest of the world. A 2021 Maryland sample (NCBI accession number ON676708) was added to this dataset for use as an outgroup.

Genomes with fewer than 180,000 non-’N’ sites were subsequently excluded. We aligned the remaining MPXV genomes against the Clade II reference genome (NC_063383) and masked the inverted terminal repeat (ITR) regions, as well as repetitive and low complexity regions, using squirrel (https://github.com/aineniamh/squirrel). As some regions of a subset of genomes were still poorly aligned, we removed stretches of sequences with many ‘N’s. Specifically, we iteratively examined each sequence using a window of size 100 nucleotides, and if the start and end of the window were each an ‘N’ and more than ≥25% of the window were ‘N’s, we masked the given window. Genomes that had either fewer than 10 (*n*=1) or more than 90 (*n*=10) substitutions relative to the reference genome were considered outliers and were excluded.

Sequences from individuals from NYC with multiple high-quality genomes were filtered to only include the earliest genome from each individual, and if there were multiple genomes with the same collection date for a given individual, the genome with the fewest ambiguities was chosen. The final dataset comprised 4044 genomes, with 757 genomes from NYC (756 generated by PHL), 1127 from the broader United States, and 2160 from the rest of the world.

The percentage of cases sequenced is based on the number of cases from OWID and the number of genomes from each region from 28 April 2022 through 28 April 2023. The number of genomes was calculated before filtering, with the exception of only counting each case from NYC with multiple genomes once. The accession IDs for all genomes used can be found in Supplementary Table S1 (all) and Supplementary Table S2 (GISAID).

### Phylogenetic analyses

We inferred a maximum likelihood tree with MAPLE^51^ using a general time-reversible (GTR) model. We extracted the B.1 clade subtree and then converted it from a bifurcating tree to a multifurcating tree using the ‘ape’ package^52^ in R v4.3.2^53^.

We performed time-calibration of the resulting multifurcating tree using a recently implemented model in BEAST v1.10^54^, which replaces the traditional likelihood with a more efficient likelihood approximation^55^ based on a Poisson model. In this approach, a starting tree scaled to substitutions/site is provided and the tree search is constrained such that only node heights and polytomy resolutions are sampled. We used a starting clock rate of 5.0x10^-5^ and a relatively uninformative gamma prior with a shape of 0.001 and scale of 1000, as well as a non-parametric coalescent tree^56^ prior with 26 evenly-placed grid-points between the most recently sampled sequence and two years prior to that time. We simulated 15 Markov chain Monte Carlo (MCMC) chains of 5x10^8^ to 1x10^9^ generations, subsampling every 25,000 iterations to continuous parameter log files and every 250,000 iterations to continuous tree files. The first 10–20% of each chain was discarded as burnin for the analyses, and the chains were then combined using LogCombiner. The resulting parameter log and tree files were resampled every 500,000 and 5,000,000 states, respectively. Model convergence and mixing was assessed in Tracer^57^, and all ESS values were >150.

To reconstruct the importation dynamics of MPXV, we used an asymmetric discrete-trait analysis (DTA) model^58^ implemented in BEAST v1.10 with samples assigned to NYC, United States (excluding NYC), and global (excluding the United States in its entirety) locales. We used the resampled empirical tree distribution described above and simulated one MCMC chain of 1x10^6^ generations, subsampling every 100 iterations to a continuous parameter log file and every 500 iterations to a continuous tree file. We discarded the first 10% of the chain as burnin and resampled the trees every 1,500 states. A maximum clade credibility (MCC) tree was then generated with TreeAnnotater 1.10 using the subsampled trees. For individual posterior tree samples, an introduction into a region is defined when a node is assigned a particular location with high probability from the posterior distribution and its parent node has a different location^59^. For the MCC tree, we define an introduction into a region when a node has a posterior probability >0.5 of the given location and its parent node has a posterior probability of ≤0.5 of the same location. To identify the cluster sizes resulting from an introduction, we performed a depth-first search starting at all internal nodes that correspond to an introduction of MPXV, traversing forward in time until a node corresponding to a different region (*i.e.*, an exportation) is encountered or there are no more nodes remaining to be explored. Transmission clusters are defined as introductions that led to at least two genomes, and introductions resulting in one genome are labeled singletons. When showcasing the time of importation for each transmission cluster, we include the entire 95% highest posterior density (HPD) intervals of the time of the most recent common ancestor (MRCA) of the transmission cluster and the time of the parent node of this MRCA.

We further explored the transmission dynamics of MPXV through the application of an episodic birth-death sampling (EBDS) model in BEAST. This phylodynamic approach enables the investigation of dynamic changes in birth, death, and sampling rates of individuals over discrete time epochs^60^. We conducted a joint phylogeny analysis, where we inferred conditionally independent trees for all clusters comprising at least 10 genomes (n=13), all derived from the same EBDS model. We implemented an EBDS model with a cutoff value of 10 months, inferring time-varying birth and sampling rates across 40 epochs, each representing one week. The model assumes a constant death rate over time, no intensive sampling events at epoch transitions, and the removal of lineages upon sampling. Gaussian Markov Random Field (GMRF) priors are employed for the birth and sampling rates to accommodate the substantial variability in effective reproduction number over time. The prior distribution for the constant death rate follows a log-normal distribution, with mean and standard deviation values estimated to yield a 95% confidence interval for infectious duration from 2 to 4 weeks. For tree inference, we applied a GTR nucleotide substitution model for all trees and a strict molecular clock model for each cluster, incorporating proper CTMC reference priors^61^ for the evolutionary rates.

We simulated 5 MCMC chains of 8x10^8^ generations each, facilitated by Hamiltonian Monte Carlo transition kernels^62^ and subsampling every 1,000 iterations to continuous parameter log files.

The effective reproduction number for each epoch is computed as

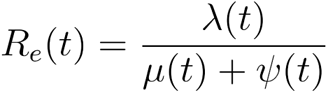

and the growth rate is calculated by

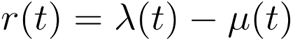

where λ, μ, and ψ are functions of time (*t*) representing the birth rate, death rate, and sampling rate, respectively.

### Demographics analysis

Correlates of membership in MPXV transmission clusters were assessed using logistic regression in R 4.0.2. Of the 756 people with a high-quality sequenced MPXV genome and in the PHL surveillance network, demographic and transmission risk metadata was available for 708 individuals. To achieve statistical convergence, we excluded 2 cis-woman and 3 individuals with unknown gender from our regression analysis, resulting in 703 included individuals.

### HIV transmission network

Because HIV has been circulating in NYC since the 1970s and we are interested in the comparison of more recent transmission clusters, we used HIV-TRAnsmission Cluster Engine (HIV-TRACE)58 to infer these clusters based on genetic distance between these virus genomes, rather than identifying clusters using phylogeographic inference. The first reported sequence for each person was aligned to the HXB2 reference PR+POL sequence (positions 2253-5094) and trimmed to PR/partial-RT to positions 2253–3749. Sequences with ≥5% ambiguous nucleotides and those ≤0.015 substitutions/site from the HXB2 reference sequence were excluded. TN93 genetic distance was compared between the first reported viral sequences from 76,910 individuals in residing in or receiving HIV care in NYC reported to NYC Department of Health and Mental Hygiene as of January 2024; an HIV transmission network was constructed using a distance threshold of 0.015 substitutions/site (resolving distances between ambiguous nucleotides for sequences with ≤1.5% ambiguities). For each clustered individual, we determined (i) if their cluster had added a case diagnosed in 2021 and (ii) if they were directly genetically linked to a virus from a case diagnosed in 2021.

Matching between individuals with an MPXV and HIV diagnosis reported to DOHMH was performed based on a 36-key deterministic matching algorithm^63^. The relationship between MPXV cases and HIV cases reported to the NYC Department of Health and Mental Hygiene was assessed using a logistic regression when expected values were ≥5 and a Fisher’s Exact Test when expected values were <5.

### Transmission lineage size distribution fitting

We fit the following distributions to the MPXV transmission cluster sizes: exponential, binomial, negative binomial, pareto, power law, and Yule-Simon. We used a minimum cluster size of two and truncated the probability distribution to the minimum cluster size. Fit was determined by the Akaike Information Criterion (AIC). Because much of the probability distribution was typically truncated during the fitting, to further examine which distributions best fit the transmission cluster sizes, we (1) subsequently simulated 10,000 transmission cluster sizes using the fitted distributions, (2) truncated the simulated data to the minimum cluster size, (3) normalized the inferred and simulated cluster sizes based on the total number of cluster sizes for each, and (4) calculated the Kullback–Leibler divergence between these two sets of normalized cluster sizes. We repeated this process for the HIV transmission cluster sizes.

### Epidemiology

To supplement the phylogenetic analyses (see above) and characterize the growth and decline of the MPXV epidemic in NYC using more traditional epidemiological approaches, we inferred the time-varying reproduction number (also known as the effective reproduction number, R_e_) and the growth rate using EpiNow2^64^, assuming a gamma-distributed generation time with a mean of 12.6 days and a standard deviation of 5.7 days and a gamma-distributed incubation time with a mean of 9.1 days and a standard deviation of 3 days. As the generation time and incubation period can vary over the course of an epidemic^33^, we also calculated R_e_ and growth rate using the approach from ref.^65^, which only uses case counts from the epidemic.

### Simulating epidemics

We simulated mpox epidemics based on the approach in ref.^13^. Briefly, this approach uses a branching process model of transmission among a men who have sex with men (MSM) population. The MSM sexual network was characterized by a Weibull distribution with parameters that were estimated by fitting a Weibull distribution to the empirical degree distribution from the Natsal dataset, which is composed of MSM who had at least one sexual partner over the course of a year. The simulations assumed a single starting case and each case transmitted the virus to their sexual partners with a constant probability per partner (*i.e.*, secondary attack rate [SAR]) over the infectious period. To determine the size of the MSM population in NYC in 2022 for the simulations, we multiplied the proportion of MSM per county in NYC based on ref.^66^ by the expected number of adult men living in each county^67^. We used the reported proportions of MSM for each county in NYC, but we used the average New York State proportion of MSM for Richmond County (Staten Island) because it was not listed in ref.^66^. This resulted in an MSM population of approximately 235,000.

We performed 1,000 simulations with this population size, SARs of 0.1 and 0.4, and infectious periods of 14 and 21 days. The effective reproduction number was calculated at each infected case as per ref.^13^. High-connectivity individuals were defined as those with a degree above a certain percentile (*e.g.*, 95th percentile, or the top 5% of connected individuals). We then calculated the proportion of high-connectivity susceptibles remaining as each case was infected.

## Data Availability

MXPV genomes from the NYC DOHMH PHL that we generated have been deposited at NCBI, and their accessions are listed in Supplementary Table S1. HIV surveillance activities are protected by New York State Redisclosure Law Articles 21 and 27-F, which prevents the submission of HIV-1 genetic sequences to public databases. Data were shared with investigators under data use agreements with UC San Diego.

## Ethical Review

This was a routine analysis of existing public health surveillance data by DOHMH surveillance analysts and thus not subject to Institutional Review Board approval at the NYC DOHMH. The IRB of the University of California at San Diego, which does not conduct public health surveillance, judged the analysis to be minimal risk [research], and “a waiver of individual authorization for the use of Protected Health Information (PHI) was granted as stipulated by the HIPAA Privacy Rule, 45 CFR 164 section 512(I).”

## Code Availability

Code is available at https://github.com/pekarj/nyc_mpox_phylogeography.

## Data Availability

All data produced in the present study are available at NCBI.

https://github.com/pekarj/nyc_mpox_phylogeography

## Acknowledgements

We thank Akira Endo for assistance in using his software to simulate mpox epidemics, Randall Collura for suggestions related to the comparison of HIV and MPXV networks, and Mark Harrington and Fabian Lim for insightful comments. We gratefully acknowledge the authors from the originating laboratories and the submitting laboratories, who generated and shared through GISAID the viral genomic sequences and metadata on which this research is based (Supplementary Table S2).

## Author contributions

J.E.P. and J.O.W. conceived the research. J.E.P., Y.W, J.C.W., Y.S., L.A.F., Y.S., P.L., M.A.S. and J.O.W. analyzed the data. J.C.W., L.A.F., H.A., T.C., F.T., and L.V.T. generated and managed the data. J.E.P. performed the simulations and wrote the first draft of the manuscript. All authors contributed to the interpretation of results and manuscript writing. J.O.W. supervised the work.

**Correspondence and requests for materials** should be addressed to J.E.P. or J.O.W.

